# Follow-up study in the ski-resort Ischgl: Antibody and T cell responses to SARS-CoV-2 persisted for up to 8 months after infection and transmission of virus was low even during the second infection wave in Austria

**DOI:** 10.1101/2021.02.19.21252089

**Authors:** Wegene Borena, Zoltán Bánki, Katie Bates, Hannes Winner, Lydia Riepler, Annika Rössler, Lisa Pipperger, Igor Theurl, Barbara Falkensammer, Hanno Ulmer, Andreas Walser, Daniel Pichler, Matthias Baumgartner, Sebastian Schönherr, Lukas Forer, Ludwig Knabl, Reinhard Würzner, Dorothee von Laer, Jörg Paetzold, Janine Kimpel

## Abstract

**Background:** In early March 2020, a SARS-CoV-2 outbreak in the ski resort Ischgl in Austria initiated the spread of SARS-CoV-2 throughout Austria and Northern Europe. In a cross-sectional study, we found that the seroprevalence in the adult population of Ischgl had reached 45% by the end of April. To answer the question of how long immunity persists and what effect this high-level immunity had on virus transmission, we performed a follow-up study in early November, 2020.

**Methods:** Of the 1259 adults that participated in the baseline study, 801 could be included in the follow-up. The study involved the analysis of binding and neutralizing antibodies and T cell responses. In addition, the incidence of SARS-CoV-2 infections in Ischgl was compared to the incidence in similar municipalities in Tyrol throughout 2020.

**Findings:** For the 801 individuals that participated in both studies, the seroprevalence declined from 51.4% (95% confidence interval (CI) 47.9 - 54.9) to 45.4% (95% CI 42.0 - 49.0). Median antibody concentrations dropped considerably but antibody avidity increased. T cell responses were analysed in 93 cases, including all 4 formerly seropositive cases that had lost antibodies in all assays, three of which still had detectable T cell memory. In addition, the incidence in the second COVID-19 wave that hit Austria in November 2020, was significantly lower in Ischgl than in comparable municipalities in Tyrol or the rest of Austria.

**Interpretation:** This study has important implications as it shows that although antibodies to SARS-CoV-2 declined, T and B cell memory can be detected for up to 8 months. Complemented by infection prevention measures a level of around 40-45% immunity in Ischgl significantly reduced local virus transmission during the second wave in Austria in November 2020.

**Funding:** Funding was provided by the government of Tyrol and the FWF Austrian Science Fund.

## Introduction

Early on in the COVID-19 pandemic, it had already become clear that individuals previously infected with the SARS-CoV-2 virus are immune to re-infection. Consequently, well-documented re-infections have been extremely rare to date, one year after the pandemic emerged ^1-3^. The question however, of how long this immunity will last, is still controversial. While some studies report a rapid loss of antibodies to SARS-CoV-2 within 2-4 months ^4,5^, especially in individuals after mild or asymptomatic infections, newer studies have shown persistence of antibodies for several months, although levels decline over time ^6^. In addition, recent studies show that T cell responses can still be detected 6-8 months after infection and most individuals after mild and asymptomatic infection retain at least T cell responses or neutralizing antibodies for up to 16-18 weeks ^7,8^. The question remains, however, if this level of immunity protects from re-infection and whether it contributes to herd immunity.

In late April 2020, we studied the seroprevalence for SARS-CoV-2 in Ischgl, a popular ski resort in the Tyrolean Alps ^9^. Ischgl was hit hard in March 2020 by the COVID-19 pandemic, and from Ischgl, the virus spread world-wide, mainly to Northern Europe and to the US ^10-12^. A total of 1473 individual, incl. 214 children, participated in our April study, corresponding to around 80% of the individuals living in Ischgl (residents and seasonal workers) at that time. We found that by the end of April 42% of the local population (45% of the adult population) had become seropositive. This was one of the highest regional seroprevalence levels reported for spring 2020 and most of these seropositive individuals had been infected in March with a drastic decline of new infections in April.

Here, we report the results of a follow-up study (Ischgl 2) performed in the first week of November 2020, among adults, 6.5 months after the first study and up to 8 months after the first infection wave in Ischgl. We found that T-cell and antibody responses could still be detected in most individuals that had recovered from infection and that very few new infections occurred from May to December, 2020.

## Material and Methods

### Study population, study design and recruitment

The ethical committee (EC) of the Medical University of Innsbruck approved the studies, baseline (Ischgl 1) with EC numbers: 1100/2020 and 1111/2020, which took place between April 21^st^ and 27^th^, 2020 and follow-up (Ischgl 2) with EC number: 1330/2020, which took place between November 2^nd^ and 8^th^, 2020. This cross-sectional epidemiological survey targeted all residents of Ischgl/Tyrol older than 18. At baseline, n=1259 adults (82.4%) of all eligible adults (n=1527) in Ischgl enrolled in the study ^9^. Of the baseline sample, n=813 (64.6%) entered the 6.5 months follow-up study. A total of n=12 of these cases were excluded from analysis due to inconsistent age (n=9), no questionnaire (n=2) and no blood sample (n=1). In total, n=91 new participants entered the study at follow up; n=12 of these were <18 years at baseline, n=1 was excluded due to missing questionnaire data. Data have been collected with Askimed, a web-based eCRF system for data collection and management.

### SARS-CoV-2 antibody tests

EDTA-plasma was analyzed for SARS-CoV-2-binding antibodies using four immunoassays. Samples were screened for anti-SARS-CoV-2-S1-protein IgA and IgG positivity by a commercially available anti-SARS-CoV-2-IgA and -IgG ELISA (Euroimmun, Lübeck, Germany), respectively, using the fully automated 4-plate benchtop instrument Immunomat™ (Virion/Serion, Würzburg, Germany). Results with respect to the obtained optical density (OD) values were interpreted according to the recommendations in the manufacturer’s information. For both assays values >1.1 were considered positive. Borderline values (0.8-1.1) in the Euroimmun IgG ELISA were rated positive, for the Euroimmun IgA ELISA borderline values were rated as negative. Additionally, each plasma was tested for anti-SARS-CoV-2-N-protein IgG (anti-N IgG) with the Abbott SARS-CoV-2 IgG immunoassay on the ARCHITECT i2000SR system (Abbott, Illinois, USA). Anti-N IgG was positive, if the obtained relative light unit (RLU) value corresponded to the manufacturer’s recommendations (>1.4). Anti-N IgG was additionally quantified using the ElecsysAnti-SARS-CoV-2 (Roche Diagnostics, Indianapolis, USA) according to manufacturer’s recommendations. The Roche assay detects antibodies against SARS-CoV-2 N protein using a double-antigen sandwich immunoassay design. This assay uses a cutoff index (COI), which is calculated using the standards provided by the manufacturer. A COI of ≥1.0 was considered positive.

### Neutralizing antibody-assay

Titers of SARS-CoV-2 neutralizing antibodies were determined using a replication defective vesicular stomatitis virus (VSV) pseudotyped with SARS-CoV-2 spike protein as described previously ^13^. Shortly, VSVΔG-GFP virus was produced on 293T cells stably expressing a C-terminally truncated version of SARS-CoV-2 spike (Wuhan variant). Four-fold serial dilutions of heat-inactivated plasma were pre-incubated with virus for 1 h at 37°C and subsequently used to infect 293T-ACE2 cells seeded the previous day. Approximately 16 h after infection, plates were analyzed in an ImmunoSpot® S5 analyzer (C.T.L. Europe, Bonn, Germany) and the number of GFP positive cells was counted. The last plasma dilution that resulted in a 50% reduction of GFP positive cells compared to virus only wells was considered as 50% neutralization titer. Titers of ≤1:4 were

### Defining seroprevalence and serostatus

Plasma samples were analyzed according to the scheme in Figure 1a. The serostatus of the samples was defined as p, d, a or n depending on the binding antibody assays:

**Figure 1.**
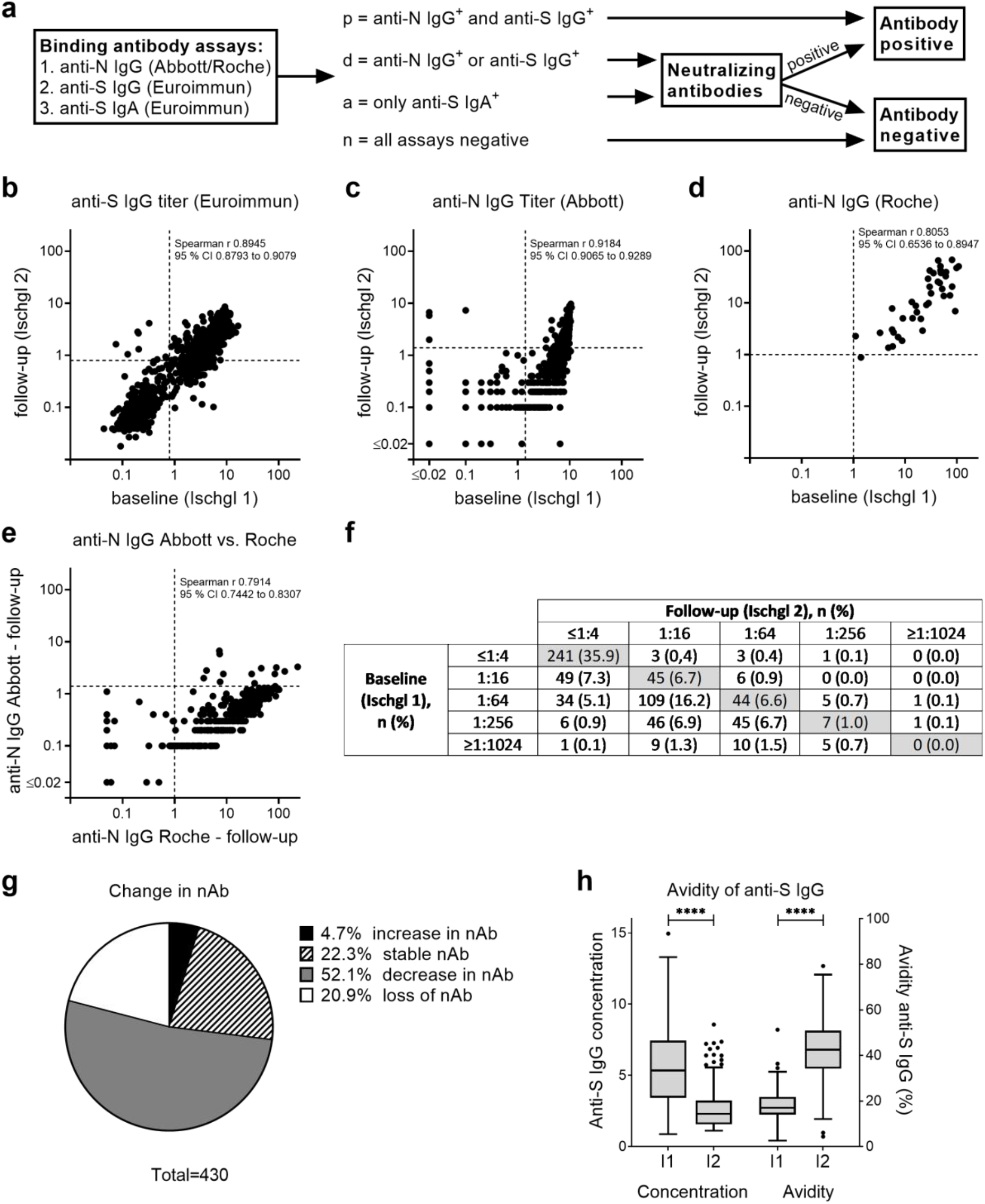
Changes in SARS-CoV-2-specific antibodies. (a) Antibodies were analyzed at baseline (Ischgl 1) and after 6.5 months follow up (Ischgl 2) using four immunoassays for binding antibodies and a neutralization assay. (b) Titers of anti-S IgG using Euroimmun ELISA. OD > 0.8 was counted as positive (dotted line), n = 801. (c) Titers of anti-N IgG using Abbott immunoassay. OD > 1.4 was counted as positive (dotted line), n = 801. (d) Titers of anti-N IgG using Roche immunoassay. OD > 1 was counted as positive (dotted line), n = 40 pairs of samples were analyzed. (e) Correlation of anti-N IgG determined via Abbott versus Roche assay at follow-up, n = 309. (b-e) Spearman r and 95% CI are depicted. (f) Titers of neutralizing antibodies. Titers ≤1:4 were counted as negative, n = 671 sample pairs were analyzed. (g) Change of neutralizing antibody titers between baseline and follow-up was analyzed for samples from f with positive (≥1:16) neutralizing antibody titers either in baseline or follow-up study. (h) Avidity of anti-S IgG was analyzed at baseline and follow-up for all samples with positive anti-S IgG titers at both time points, n = 218. Temporal trend in the median (IQR) concentration of anti-SARS-CoV-2 IgG antibodies as compared to the median binding affinities (avidities) of the antibodies (%). The lower and upper bars represent the minimum and maximum values, respectively. Dots represent outliers. Statistics were determined using Students t-test for paired samples, ^****^ p<0.0001.

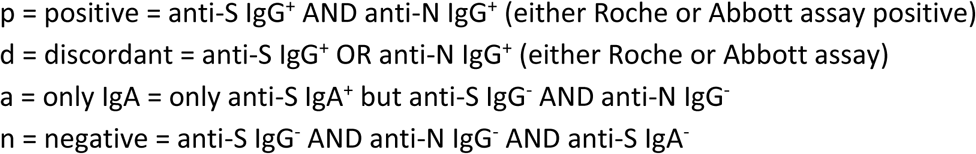

To calculate the seroprevalence, all individuals with serostatus p were considered as seropositive. For serostatus d and a individuals were considered as seropositive when they had neutralizing antibodies ≥1:16.

### Avidity of SARS-CoV-2-specific anti-S IgG

A kit was used according to manufactures instructions to determine of the avidity of anti-S antibodies (Euroimmun AG, Lübeck, Germany, REF: EI 2606-9601 G). Shortly, plasma samples, in a 1:101 dilution, were added to microplate wells coated with an S1-domain of the spike protein of SARS-CoV-2 and incubated in duplicates for 60 minutes at 37°C. After three washing steps, the pairs of samples were treated either with 200 µl urea or 200 µl PBS and were further incubated for 10 minutes. Plates were washed again. For the detection of remaining antibody, 100 µl of peroxidase-conjugated anti-human IgG were added, incubated for another 30 minutes at 37°C and plates were subsequently washed. For the colorimetric signal detection TMB substrate was used. Reaction was stopped after a 30-minute incubation at room temperature and plates were measured at wavelengths of 450 nm (signal) and 620 nm (reference wavelength for background subtraction) using a Tecan Sunrise Reader (Grödig, Austria). The relative avidity index (RAI) for each sample corresponded the ratio of the optical densities (ODs) with and without urea incubation.

### Analysis of SARS-CoV-2-specific T cell responses

For details regarding T cell isolation and analysis see supplementary methods.

To study SARS-CoV-2 specific T cell responses Prot_S, Prot_M and Prot_N SARS-CoV-2 PepTivator® (Miltenyi Biotech, Bergisch-Gladbach, Germany) peptide pools consisting mainly of 15-mer sequences with 11 amino acids overlap covering the immunodominant sequence domains of the surface (or spike) glycoprotein (pepS), and the complete sequence of the membrane glycoprotein (pepM) as well as the nucleocapsid phosphoprotein (pepN) were used. As positive control PepTivator® CEF MHC Class 1 Plus (pepCEF) was used, consisting of 32 MHC class 1-specific peptides of 8-12 amino acids in length derived from human cytomegalovirus (HCMV), Epstein-Barr virus (EBV), and influenza A virus. Alternatively, cells were stimulated with Phytohaemagglutinin (PHA, Sigma) at a concentration of 10 µg/ml.

To expand SARS-CoV-2 reactive T cells, 2×10^6^ PBMCs in 500 μL of RPMI medium supplemented with 2% human AB serum were stimulated with 1 µg/ml pepS, pepM or pepN peptide pools in the presence of 20 U/ml interleukine-2 (IL-2). As negative control, cells were cultured with IL-2 alone. PepTivator pepCEF at 1 µg/ml in the presence of IL-2 was used as positive control. At day 3, 500 μL fresh RPMI medium supplemented with 2% human AB serum and 40 U/ml IL-2 was added. At day 7, cells were harvested and counted. A total of 1×10^5^ cells were re-stimulated with or without 1 µg/ml pepS, pepM or pepN peptide pools. As positive controls, cells were stimulated with 10 µg/ml PHA or re-stimulated with PepTivator pepCEF at 1 µg/ml. Specific T cell responses were analyzed in an IFNγ enzyme-linked immunospot (ELISPOT) or by intracellular (IC) cytokine and cell surface staining (Supplementary Methods).

### Statistical analysis

Statistical analyses were performed in Stata 16.1 (StataCorp. 2019. Stata Statistical Software: Release 16. College Station, TX: StataCorp LLC) and GraphPad Prism version 8.2.0 and 9 (GraphPad Software, Inc., La Jolla, CA, USA). Demographic characteristics were tabulated using descriptive statistics including the calculation of means ± standard deviations (or median and interquartile range (IQR)) for continuous measures and numbers (%) for categorical measures. 95% confidence intervals for binomial proportions, including crude prevalence estimates of seroprevalence, were calculated using the Clopper-Pearson estimation method. Cross-sectional seroprevalence was calculated in November for the whole sample, seroprevalence at baseline and follow-up was also calculated for individuals for whom data was available for both.

Quantitative variables were compared across groups using parametric (ANOVA) and non-parametric methods (Mann-Whitney U-test, Kruskal-Wallis test). Differences in quantitative variables at baseline and follow-up were compared using parametric (Student’s t-test for paired samples, repeated measures ANOVA) and non-parametric (Wilcoxon signed-rank test) tests. Associations between categorical variables were tested using χ^2^ test, with Fisher’s exact test where appropriate. A two-tailed value of p <0.05 was considered statistically significant for all comparisons.

### SARS-CoV-2 transmission in Ischgl in 2020

We compared daily numbers of confirmed SARS-CoV-2 PCR-positive cases of Ischgl with 13 control municipalities. We selected 5% of all municipalities in the federal state of Tyrol (279 in total) that are most similar to Ischgl in terms of the Mahalanobis distance in covariate space. The covariates used to calculate the Mahalanobis distance were population size, settlement area per square km, share of female population, share of people under 16 years, share of people older than 65 years, number of commuters, share of people with tertiary education, and hotel bed capacity. The matching approach selected control group municipalities which are most similar to Ischgl. The selected control municipalities were Eben am Achensee, Ellmau, Fiss, Gerlos, Lermoos, Leutasch, Mayrhofen, Nauders, Neustift im Stubaital, Seefeld in Tirol, Serfaus, Sölden, Tux. These are all very tourism-intense holiday towns in Tyrol, and very comparable to Ischgl in terms of the covariates used. More details on the calculation of the Mahalanobis distance are available upon request. All calculations were executed with the software R (version 4.03), and the R-package Synth (8).

To test whether the trends are significantly different between Ischgl and the 13 control municipalities during the second wave, we estimated an event-study model. In particular, we divide our daily data into weekly periods starting from September 1st and calculated for each week the Difference-in-Difference in the 7-day moving average of new cases (per 100,000 inhabitants) between the group of control municipalities and Ischgl. The regression equation (1) is given by:

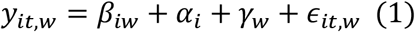

*y*_*it,w*_ denotes the 7-day moving average of new cases in municipality *i* (Ischgl or the average control municipality), at day *t* and week *w*. α_*i*_ are municipality-fixed effects, γ_*w*_ denote week fixed effects, and ε is the remainder error term. Our coefficient of interest is *β*_*iw*_ and measures the difference in the outcome variable between Ischgl and the control municipalities between week *w* and the reference period. As reference period we chose the second week of September, which was several weeks before the second wave hit Austria. Our sample ends at December 14 and, therefore, included 16 weeks. Thus, we estimated 16 *β*_*iw*_*’s*, which are plotted in a graph along with the corresponding 95% confidence intervals (Figure 3b)

## Results

### Study population

In the first week of November 2020, 6.5 months after our first seroprevalence study in the ski-resort Ischgl (baseline study = Ischgl 1), we again invited all adult Ischgl residents to participate in a study on the level of local immunity to SARS-CoV-2 (follow-up study = Ischgl 2). In April, an estimated 1527 adults lived in Ischgl, of these 1259 participated in Ischgl 1 (82.4%). In November, an estimated 1304 adults were living in Ischgl, this decrease reflects the absence of seasonal workers. Of these, 904 adults participated in November (69.3% participation rate). Of the 1259 study participants at baseline, 801 individuals participated again in November and were included in the analysis in the follow-up study presented here (63.6%, Supplementary Figure 1). Age distribution was similar in the adult population in both studies, while there was a slight bias towards female participants in Ischgl 2 (54.9%) relative to Ischgl 1 (51.9%) (Supplementary Table 1).

### Antibodies to SARS-CoV-2 were still detectable although the levels declined

The testing strategy for antibodies to SARS-CoV-2 is depicted in Figure 1a. IgG antibodies against spike (S) and nucleoprotein (N) were determined using commercials available immunoassays, anti-SARS-CoV-2-IgG ELISA (Euroimmun, Lübeck, Germany) for anti-S IgG, Abbott SARS-CoV-2 IgG immunoassay (Abbott, Illinois, USA) and ElecsysAnti-SARS-CoV-2 (Roche Diagnostics, Indianapolis, USA) for anti-N IgG. We observed that many individuals had lost anti-N IgG antibodies in the Abbott anti-N IgG-Abbott test in Ischgl 2, but turned out to still have anti-N IgG antibodies in the Roche anti-N IgG-Roche assay (Supplementary Table 2). For individuals negative in all three assays, anti-S IgA antibodies were determined using an anti-SARS-CoV-2-IgA ELISA (Euroimmun, Lübeck, Germany). We defined the serostatus groups p (positive), d (discordant), a (IgA to S protein only), and n (negative in all test), based on the outcome of the commercial antibody assays as explained in Material and Methods and in Figure 1a.

For all individuals with serostatus d and a neutralizing antibodies were determined. The following individuals were considered seropositive: p (anti-S IgG and anti-N IgG positive), d (discordant) plus neutralizing antibody positive; a (anti-S IgA only) plus neutralizing antibody positive. We have previously described a seroprevalence of 45.0% (95% CI 42.2 – 47.8) for all adult participants at Ischgl 1 (n=1259) ^9^. For the follow-up study, there was a slight bias for participation of baseline study seropositive individuals, while fewer negative individuals returned. This resulted in a baseline study seroprevalence of the subpopulation included in both studies (n=801) of 51.4% (95% CI 47.9 – 54.9). Here, we determined the seroprevalence for all adult participants at Ischgl 2 (n=891) with 44.7% (95% CI 41.4 to 48.0). For individuals that participated in both studies (n=801), the level of seropositivity declined between both studies to 45.4% (95% CI 42.0 - 49.0) in Ischgl 2 (Supplementary Table 3). From the 90 individuals that only participated in Ischgl 2 and not in Ischgl 1, 37.8% (95% CI 27.8 - 48.6) were seropositive in Ischgl 2 (Supplementary Table 3). The individuals that only participated in Ischgl 2 were not otherwise included in the following analysis.

Median antibody levels to SARS-CoV-2 significantly declined in each assay - by 50% in the anti-S IgG assay (*n* = 801, Z =22.0, p< 0.001), 88.9% in the anti-N-IgG-Abbott (*n* = 801, Z = 19.4, p< 0.001) and 58.6% in the anti-N-IgG-Roche assay (*n* = 40, Z = −4.4, p< 0.001) (Figure 1 b-d and Supplementary Table 2). In several individuals, antibodies were lost for one of the antigens switching from positive “p” to discordant “d” (p-d) (Table 1). Only 4 of 364 (1.1%, 95% CI 0.3-2.8) individuals with serostatus p (positive for anti-S and anti-N antibodies) in Ischgl 1 were tested negative in all commercial antibody test kits in Ischgl 2 (group p-n, Table 1). Interestingly, 3 of the 4 individuals in the p-n group (75%) still retained neutralizing antibodies (Table 1). Of the 8 sera that were only positive for IgA antibodies to S protein in Ischgl 2, 2 had neutralizing antibodies (Table 1).

**Table 1.** Serostatus at baseline (Ischgl 1) and follow-up (Ischgl 2)

|  |  | Serostatus at follow-up (Ischgl 2), n (%), NT<sup>+</sup>/total NT assays |  |  |  |  |
| --- | --- | --- | --- | --- | --- | --- |
|  |  | Positive - p | Discordant - d | IgA only - a | Negative - n | Total |
| **Serostatus at baseline (Ischgl 1), n (%), NT<sup>+</sup>/total NT assays** | Positive - p | 276 (34.5)<br>244 NT<sup>+</sup>/276 | 82 (10.2)<br>63 NT<sup>+</sup>/82 | 2 (0.2)<br>2 NT<sup>+</sup>/2 | 4 (0.5)<br>3 NT<sup>+</sup>/4 | 364 |
|  | Discordant - d | 4 (0.5)<br>2 NT<sup>+</sup>/4 | 30 (3.7)<br>13 NT<sup>+</sup>/30 | 1 (0.1)<br>0 NT<sup>+</sup>/1 | 8 (1.0)<br>2 NT<sup>+</sup>/7 | 43 |
|  | IgA only - a | 0 (0.0) | 1 (0.1)<br>0 NT<sup>+</sup>/1 | 3 (0.4)<br>0 NT<sup>+</sup>/3 | 7 (0.9)<br>0 NT<sup>+</sup>/7 | 11 |
|  | Negative - n | 4 (0.5)<br>3 NT<sup>+</sup>/4 | 5 (0.6)<br>2 NT<sup>+</sup>/5 | 2 (0.2)<br>0 NT<sup>+</sup>/2 | 372 (46.4)<br>6 NT<sup>+</sup>/246 | 383 |
|  | Total | 284 (35.5) | 118 (14.7) | 8 (1.0) | 391 (48.8) | 801 |

NT<sup>+</sup> = titer ≥1:16

Also the neutralizing antibody titers declined in most individuals that were neutralizing antibodies positive in Ischgl 1, and 90 of 430 (20.9%, 95% CI 17.2 to 25.1) individuals completely lost neutralization capacity in our assay (Figure 1f, g). Interestingly, of these 90 individuals that lost neutralizing antibodies in Ischgl 2, 1 had IgA to S, 33 had IgG antibodies to either N or S and 32 had antibodies to both antigens in the commercial antibody assays.

We then studied the avidity of the antibodies in individuals that were anti-S IgG positive in Ischgl 1 and 2 (Figure 1h). While the antibody titers to S had declined over time, the avidity significantly increased as a sign of antibody maturation. Thus, despite the lower concentration of antibodies in Ischgl 2, the antibodies that persisted show a higher binding strength and thereby most likely an improved functionality.

### T cell response

In Ischgl 1, T cell response had not been analysed. To study if the decline in humoral immunity was compensated at least partially by T cell responses in Ischgl 2 participants, we analyzed the response to peptide pools derived from S, M and N protein of SARS-CoV-2 by ELISPOT (Figure 2a, b) and intracellular cytokine staining (ICS) analysis (Figure 2c, d). We analyzed a total of 71 antibody positive individuals from Ischgl 1 who remained positive (p-p, n=28, all NT^pos^), became discordant (p-d, n=38, 29 NT^pos^/9 NT^neg^), or IgA only (p-a, n=1, NT^pos^) or negative (p-n, n=4, 3 NT^pos^/1 NT^neg^) in Ischgl 2. Individuals that were antibody negative in both studies (n=22, all NT^neg^) served as a control group. As positive controls, we used pepCEF and PHA, and T cell response to these stimulations were comparable in all investigated groups (Supplementary Figure 2).

**Figure 2.**
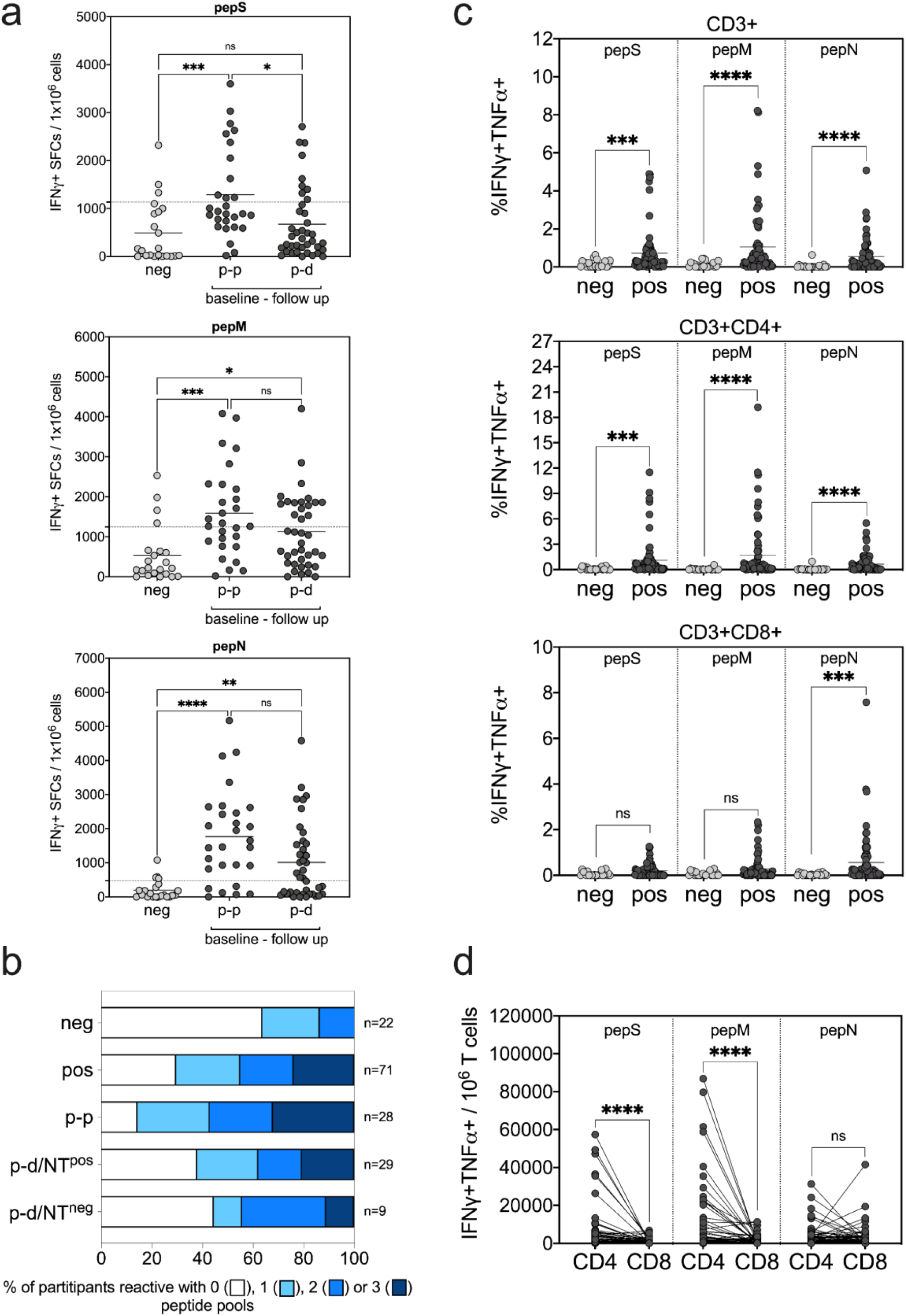
T cell responses against SARS-CoV-2-derived peptide pools. Analysis of SARS-CoV-2-specific T cells by (a,b) IFNγ ELISPOT assay and (c,d) IFNγ/TNFα intracellular cytokine staining (ICS) after a 7-day in vitro expansion followed by re-stimulation with pepS, pepM and pepN peptide pools. Values after peptide re-stimulation were normalized to non-stimulated samples in both ELISPOT and ICS. (a) IFNγ positive SFCs per 10^6^ cells are shown after pepS, pepM and pepN re-stimulation. Negative study group (neg) is compared to baseline positive with a follow up serostatus of positive (p-p) and discordant (p-d). Dotted lines show the cut-off of the assay, above which T cell responses were declared reactive in the ELISPOT assay and are defined as the mean + 1×SD of the negative group for the respective peptide pool. (b) Proportion of participants reactive with 0, 1, 2 or 3 peptide pools is shown in baseline antibody negative and baseline antibody positive study groups. Baseline antibody positive participants were additionally divided according to serostatus in follow-up study, p-p (baseline and follow-up positive), p-d/NT^pos^ (baseline positive, follow-up discordant but neutralizing antibody positive) and p-d/NT^neg^ (baseline positive, follow-up discordant but neutralizing antibody negative). (c) Percentage of IFNγ/TNFα-producing CD3+ total T cells, CD3+CD4+ helper T cells and CD3+CD8+ cytotoxic T cells after peptide re-stimulation are shown. After ICS living/singlet cells were gated for CD3+ total, CD3+CD4+ and CD3+CD8+ T cells and the indicated percentage depicts the frequency of IFNγ/TNFα double-positive cells in the respective population of the sample stimulated with the test peptide minus the frequency of the non-stimulated control. (d) Number of IFNγ/TNFα double-positive CD4+ and CD8+ cells per 10^6^ CD3+ total T cells were calculated and depicted. Statistical analysis for data in (a) were done by ANOVA and significance were calculated by Kruskal-Wallis test followed by Dunn’s multiple comparison, for data (c) and (d) significance were calculated by Mann-Whitney test (ns, not significant; ^*, **, ***^ and ^****^ represent p values <0.05, <0.01,<0.001 and <0.0001, respectively).

Although approximately one third of individuals in the seronegative group showed T cell responses to at least one peptide pool (Table 2, Figure 2b), the responses were significantly higher in baseline seropositive individuals (Figure 2a, b, c). The responses to pepM and pepN pools were not significantly different between the different groups of positive individuals studied. Stimulation with pepS pool resulted in no significant difference in responses between n-n and p-d groups, however the response in p-p group was found significantly higher compared to both n-n and p-d groups. The T cell response was primary elicited by CD4+ T cells and to a lesser extent by CD8+ T cells for the S and M protein peptide pools, while CD4+ and CD8+ T cells contributed equally to the response against the N peptide pool (Figure 2c, d).

**Table 2.** Number of T cell reactive* and neutralizing study participants (n=93)

| Number of<br>reactive peptide<br>pools | baseline<br>neg | baseline<br>pos | baseline - follow up |  |  |  |  |  |
| --- | --- | --- | --- | --- | --- | --- | --- | --- |
|  |  |  | p-p<sup>§</sup> | p-d<br>(NT<sup>pos</sup>) | p-d<br>(NT<sup>neg</sup>) | p-a<sup>§</sup> | p-n<br>(NT<sup>pos</sup>) | p-n<br>(NT<sup>neg</sup>) |
| 0 | 14/22<br>(63.6)<sup>#</sup> | 21/71<br>(29.6) | 4/28<br>(14.3) | 11/29<br>(37.9) | 4/9<br>(44.4) | 0/1<br>(0) | 1/3<br>(33.3) | 1/1<br>(100) |
| 1 | 5/22<br>(22.7) | 18/71<br>(25.4) | 8/28<br>(28.6) | 7/29<br>(24.1) | 1/9<br>(11.1) | 0/1<br>(0) | 2/3<br>(66.6) | 0/1<br>(0) |
| 2 | 3/22<br>(13.6) | 15/71<br>(21.1) | 7/28<br>(25.0) | 5/29<br>(17.2) | 3/9<br>(33.3) | 0/1<br>(0) | 0/3<br>(0) | 0/1<br>(0) |
| 3 | 0/24<br>(0) | 17/71<br>(23.9) | 9/28<br>(32.1) | 6/29<br>(20.7) | 1/9<br>(11.1) | 1/1<br>(100) | 0/3<br>(0) | 0/1<br>(0) |

\* IFNγ + SFCs / 10<sup>6</sup> cells > mean + 1 × SD of negative study participants; <sup>#</sup> number of reactive samples/number of total samples (% reactive); <sup>§</sup> all NT<sup>pos</sup> at follow-up; baseline – follow up serostatus p-p: positive - positive, p-d: positive - discordant, p-a: positive - IgA only, p-n: positive – negative, NT<sup>pos</sup>: neutralizing antibody positive (titer ≥ 1:16) at follow-up; NT<sup>neg</sup>: neutralizing antibody negative (titer ≤ 1:4) at follow-up

When analyzing the number of reactive peptide pools, we found significant difference between baseline antibody negative and positive study groups (Table 2, p=0.007). When comparing baseline negative participants with baseline positives in the different subsets in the follow-up, the number of reactive peptide pools in the p-p group but not p-d/NT^pos^ and p-d/NT^neg^ groups was significantly higher than in the n-n group (p-p versus n-n p=0.001, p-d/NT^pos^ versus n-n p=0.091 and p-d/NT^neg^ versus n-n p=0.261). Thus, T cell response in term of the number of reactive peptide pools correlated with declining serostatus with p-p > p-d/NT^pos^ > p-d/NT^neg^ (Table 2, Figure 2b). Importantly, we found that of the 9 individuals that had lost binding antibodies partially and neutralizing antibodies completely (p-d/NT^neg^), 5 still had a detectable T cell response, thus those individuals could potentially still have some T cell-mediated immune protection. Of the four individuals in the p-n group that had lost all antibodies in the commercial assays, 3 still retained neutralizing antibodies, two of which hat detectable T cell responses and only one individual had lost all binding and neutralizing antibodies and also showed no T cell responses (Table 2).

### Virus transmission in Ischgl

Between Ischgl 1 and 2, 4 individuals seroconverted, of which one individual had been tested positive by PCR in October 2020. Two of the remaining 3 reported symptoms compatible with COVID-19 between the two studies (Supplementary Table 4).

We then investigated whether the high level of immunity in Ischgl, which was still detected in November, had limited virus transmission during the second wave of SARS-CoV-2, which hit Austria in autumn 2020. We compared daily numbers of confirmed SARS-CoV-2 PCR-positive cases of Ischgl with 13 control municipalities selected from Tyrol. Figure 3a shows the 7-day moving average of daily new confirmed COVID-19 cases per 100,000 inhabitants for Ischgl and the control municipalities. Three things stick out: First, the figure confirms that Ischgl was severely hit during the first wave of the pandemic, with the daily number of cases being an order of magnitude larger compared to the control municipalities. Second, during the summer in both Ischgl and the control municipalities the number of cases converged to (almost) zero, as in most places in Europe. Third, with the start of the second wave in October 2020, the trend between the two groups sharply diverged, with the control municipalities showing a substantially larger number of confirmed cases (Figure 3a).

**Figure 3.**
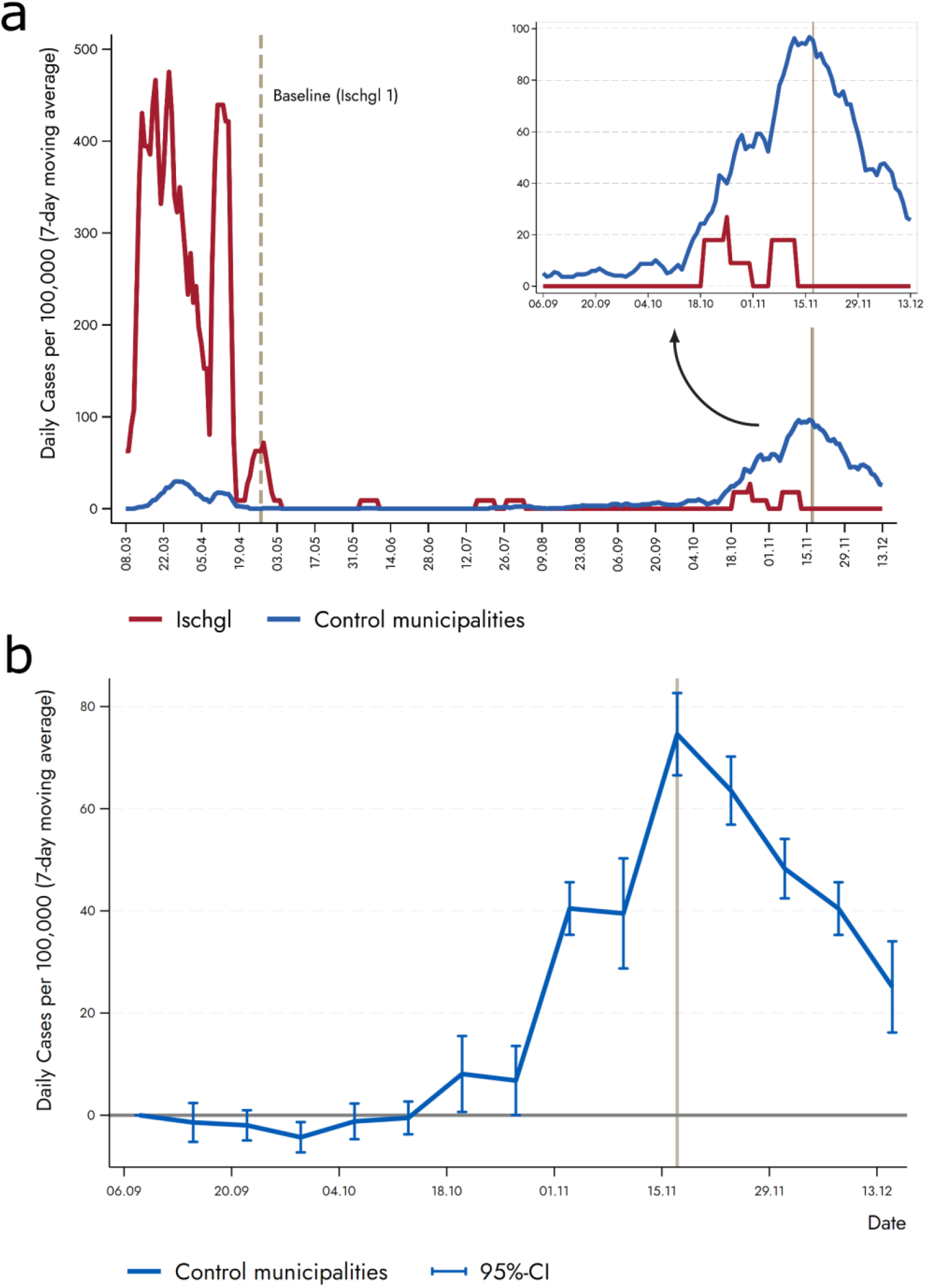
Lower incidence of new infections in Ischgl compared to low-prevalence villages. (a) Vertical solid line represents the second countrywide lockdown in autumn 2020, which took place in November 17 (a first and lighter lockdown took place on November 2). The control municipalities are Eben am Achensee, Ellmau, Fiss, Gerlos, Lermoos, Leutasch, Mayrhofen, Nauders, Neustift im Stubaital, Seefeld in Tirol, Serfaus, Sölden, and Tux. (b) The figure displays results from regression equation (1). The plotted coefficients represent the weekly difference in the 7-day moving average of new cases between Ischgl and the control municipalities relative to the reference period (2^nd^ week of September). The coefficient for each week is shown together with a 95%-confidence interval. The vertical solid line shows the second (strict) lockdown in Austria.

Figure 3b reports the results of our event study. The plotted coefficients display the weekly difference in the 7-day moving average of new cases between Ischgl and the control municipalities relative to the reference period. It shows that at the beginning of September, the difference between the two groups of municipalities was basically zero. By the end of October, the number of new cases in the control municipalities started to significantly increase relative to Ischgl. We continued to find statistically different coefficients up until Christmas, the end of our observation period (Figure 3b). This difference supports the hypothesis that the high seroprevalence level of about 40-45% found in Ischgl in November, limited viral transmission.

## Discussion

We studied immunity to SARS-CoV-2 in the ski-resort Ischgl, one of the hotspots during the first wave of the pandemic in March, 2020. We performed antibody testing of a large share of the Ischgl population in April 2020 and again 6.5 months later, in November 2020. Longitudinal data allowed us to track the level of antibodies for 801 participants (63.6% of the baseline adult study population) over time. We show, that antibody levels declined within the first 7-8 months after infection, but that the binding capacity of antibodies increased and antibodies and/or T cell responses were still detectable in all but one individual.

Our general observation is typical for viral infections, where within a few weeks after the infection, antibody levels peak and then slowly decline to a lower level that persists for longer periods, depending on the virus ^14^. This has also been shown previously for the infection with SARS-CoV-2 ^6^. However, the question of how long immunity lasts for SARS-CoV-2 has not been fully resolved.

Immunity to the human coronaviruses that cause a common cold only lasts for 1 to 2 years and repeated infections with the same virus are common. However, as also suspected to occur for SARS-CoV-2, reinfection with human coronaviruses may also be the result of viral immune escape ^15^. However, SARS-CoV-2 causes a more severe, systemic infection, where immunity may last longer. Several authors have detected antibodies to SARS-CoV-2 for several months after infection ^16^. This notion is supported by other studies reporting that immune response to SARS-CoV-1 (an infection associated with severe illness) remains detectable for longer than 17 years ^17,18^.

Several newer publications have found, similar to our study, but for a shorter average observation period, that immunity as defined by T cell and/or neutralizing antibody response lasts for up to several months and that T cell and antibody responses can be discordant ^8,19^. A more recent study, that has analyzed the T and B cell responses in great detail and in some patients for up to 8 months, but on average for a shorter period, concluded that immune memory is indeed preserved, in accordance with our findings ^7^.

The majority of the 412 adults that were seropositive during the Ischgl 1 study in late April and analyzed again in early November, Ischgl 2, had been infected already in March during the first wave. Thus, this study, to our knowledge, represents one of the longest and largest follow-up studies published so far. The conclusion that immunity may last for at least up to 8 months is in accordance with the fact that reinfections worldwide are still rare, although increasingly reported in the literature ^1-3,20,21^. However, none have been observed in Ischgl. Furthermore, this study, in contrast to similar studies, was performed in a population in a defined area and the transmission within this population could be observed in parallel with the study of immunity in the laboratory. We found that the incidence of SARS-CoV-2 in Ischgl was significantly lower than in comparable municipalities during the second wave that hit Austria in November. We had already argued in our first study, that with a seroprevalence of 40-45% and in the presence of mild non-pharmacological interventions, Ischgl may have been near the herd immunity threshold, as the incidence of SARS-CoV-2 infections had declined drastically in April. This is in accordance with a second mathematical modelling study showing that herd immunity can be achieved at a lower rate depending on population heterogeneity and social activity ^22^. However, also in other parts of Austria, the incidence declined in April, which made it difficult to draw firm conclusions. However, when the second wave hit Austria 8 months later, the reduced incidence in Ischgl relative to comparable municipalities (as well as to the rest of Austria) strongly indicates that indeed the level of immunity to SARS-CoV-2 helped to curb new infections in Ischgl. Given the fact that there was a high level of seroprevalence still present among the population of Ischgl in November, the ‘lock-down light’ measures were sufficient to effectively suppress local virus transmission, which was not the case in the rest of Austria (https://www.euractiv.com/section/coronavirus/news/austria-to-enter-partial-november-lockdown/). In this respect it is important to note that non-pharmacological interventions (NPI) have been identical for Ischgl and the control municipalities since April.

Our findings suggest that seropositivity levels of around 40-45% might suffice to stop the virus when some social distancing and NPIs are in place. Now that vaccines start to roll-out in Europe, the case of Ischgl indicates that virus transmission can be successfully controlled if social distancing measures are continued, at levels of vaccine coverage below the predicted 70 to 80% herd immunity threshold.

## Data Availability

Upon publication of our study, the data and codes will be available from the corresponding authors upon receipt of a suitable request.

## Acknowledgments

We thank Bianca Neurauter for excellent assistance when organizing the study. We thank Brigitte Müllauer, Lisa-Maria Raschbichler, Albert Falch, Maria Huber, Teresa Harthaller, and Eva Hochmuth for excellent technical support. We thank the AGES and Franz Allerberger for providing SARS-CoV-2 qPCR data and critical discussion. We thank Peter Felsner (BD Biosciences) and Csaba Merschdorf and Attila L. Edelmann (CTL Europe GmbH, Bonn, Germany) for their support on flow cytometry and ELISPOT analysis.

## Conflict of interest

No conflicts of interests exist. The funders had no role in the design of the study; in the collection, analyses, or interpretation of data; in the writing of the manuscript, or in the decision to publish the results.

## Funding

The study has been supported by the state of Tyrol. KB has been supported by a FWF Austrian Science Fund Lise Meitner Award [M-3069-B].

## Notes

### Competing Interest Statement

The authors have declared no competing interest.

### Author Declarations

Ethical committee of the Medical University of Innsbruck

