## Supplementary Materials for "Follow-up study in the ski-resort Ischgl: Antibody and T cell responses to SARS-CoV-2 persisted for up to 8 months after infection and transmission of virus was low even during the second infection wave in Austria"

**Supplementary material**

| LIST OF VOLUNTARY STUDY MEMBERS, ALL FROM AUSTRIA | |
|---|---|
| Agnes Scharrer | Liesa-Marie Schreiber |
| Albert Falch | Lisa-Maria Raschbichler |
| Alina Mracsna | Magdalena Fruhwirth |
| Anastasia von Canal | Maria Holzknecht |
| Anna Haslwanter | Maria Huber |
| Benedikt Leonhard | Martin Anegg |
| Bianca Neurauter | Martin Klieber |
| Brigitte Müllauer | Mathias Baumgartner |
| Carina Praxmarer | Michael Unterhofer |
| David Bante | Nina Fernbach |
| Eva Hochmuth | Rosmarie Gstraunthaler |
| Fatima Aslam | Sophia Schmidt |
| Frederik Radvan | Teresa Harthaller |
| Hanna Salvotti | Theodora Todorovic |
| Jonas Huber | Theresa Genser |
| Katharina Wagner | Valentin Schiessendoppler |
| Lena Hinterstoisser | Verena Söll |
| Leonie Mauser | |

**Supplementary methods**

Peripheral blood mononuclear cell (PBMC) isolation

40 ml of whole blood was collected in 9 ml S-Monovette® EDTA tubes (Sarstedt, Nümbrecht, Gemany). PBMCs were isolated by layering 20 ml EDTA blood on 15 ml Pancoll human (PAN-Biotech, Aidenbach, Germany) and subsequent density-gradient centrifugation for 30 minutes at 1800 rpm. PBMCs were collected from the interphase, washed twice in PBS and counted. PBMCs were cryopreserved in heat-inactivated fetal calf serum (FCS, Gibco) supplemented with 10% DMSO (Gibco) and stored in liquid nitrogen until use.

Interferon-γ (IFNγ) ELISPOT assay following 7-day *in vitro* expansion

Following 7-day *in vitro* expansion, cells were harvested, counted and analyzed in an IFNγ enzyme-linked immunospot (ELISPOT) assay (ImmunoSpot®, C.T.L. Europe, Bonn, Germany) according to the manufacturer's protocol. Briefly, a total of $1\times10^5$ cells per well were placed in anti-human IFNγ mAb-coated plates in 100 μL RPMI medium supplemented with 2% human AB serum and were re-stimulated with or without 1 μg/ml pepS, pepM or pepN peptide pools. As positive controls, cells were stimulated with 10 μg/ml PHA or re-stimulated with PepTivator pepCEF at 1 μg/ml. After 24 h incubation, IFNγ-producing cells were visualized by staining with anti-human IFNγ-FITC, FITC-HRP and substrate solution according to the manufacturer's protocol. Spots were counted using an ImmunoSpot® S5 analyzer (C.T.L. Europe, Bonn, Germany). IFNγ-producing spot forming cells (SFC) per $1\times10^6$ cells corresponding to specific T cell responses were calculated from the number of SFCs after specific stimulation minus SFCs of negative control without stimulation. Stimulations were considered reactive when spot count was higher than the mean of the spot count plus 1×SD of the study negative group after the respective stimulation.

Intracellular cytokine staining

SARS-CoV-2 peptide-specific T cells were further characterized by intracellular (IC) cytokine and cell surface staining. For intracellular cytokine and cell surface staining after 7-days *in vitro* expansion, $1\times10^5$ cells per well in 100 μL RPMI medium supplemented with 2% human AB serum were re-stimulated with or without 1 μg/ml pepS, pepM or pepN peptide pools. As positive controls, cells were either stimulated with 10 μg/ml PHA or re-stimulated with PepTivator pepCEF at 1 μg/ml. After 3.5 hours, BD GolgiPlug™ protein transport inhibitor (BD Austria, Vienna) was added according to the manufacturer's instructions. After an 18-hour incubation, surface staining was performed with PE-Cy7 anti-human CD3 (clone SK7), BV510 anti-human CD4 (clone SK3) and BV421 anti-human CD8 (clone RPA-T8) monoclonal antibodies all purchased from BD (BD Austria, Vienna). For live/dead staining BD Horizon™ Fixable Viability Stain 780 (BD Austria, Vienna) was used according to the manufacturer's instructions. IC staining was performed using BD Cytofix/Cytoperm™ fixation/permeabilization kit (BD Austria, Vienna) according to the manufacturer's instructions using BD FastImmune™ PE anti-human IFNγ (clone 25723.11) and APC anti-human TNFα (clone 6401.1111) antibodies (BD Austria, Vienna). All samples were measured on a BD FACSCanto™ II cytometer (BD) and data were analyzed using FlowJo v10 software (BD).

**Supplementary Table 1. Study population**

| | Baseline (Adults)* | Baseline Analysis Sample (Adults)** | Lost to Follow-up[#] | New at Follow-up[$] | Excluded (Follow-up sample) | Total Follow-up | T Cell Sample[§] |
|---|---|---|---|---|---|---|---|
| **Number of participants, n (%)** | | | | | | | |
| **Male** | 604 (48.0) | 361 (45.1) | 238 (53.4) | 52 (57.8) | 5 (41.7) | 413 (46.4) | 48 (51.6) |
| **Female** | 653 (51.9) | 440 (54.9) | 206 (46.2) | 38 (42.2) | 7 (58.3) | 478 (53.7) | 45 (48.4) |
| **Diverse** | 2 (0.2) | 0 (0.0) | 2 (0.5) | 0 (0.0) | 0 (0.0) | 0 (0.0) | 0 (0.0) |
| **Age (years), mean (SD), median** | | | | | | | |
| **Male** | 44.9 (16.2) 44.0 | 45.5 (15.6) 46.0 | 43.7 (16.9) 41.0 | 43.4 (19.7) 41.5 | 54.2 (29.4) 43.0 | 45.2 (16.3) 45.0 | 45.1 (12.5) 47.0 |
| **Female** | 45.6 (16.7) 45.0 | 45.7 (16.0) 45.0 | 45.6 (17.9) 43.5 | 39.7 (18.4) 35.0 | 44.6 (24.1) 40.0 | 45.2 (16.1) 45.0 | 42.8 (13.8) 42.0 |
| **Adults (≥ 18)** | 45.2 (16.5) 44.0 | 45.6 (15.8) 45.0 | 44.5 (17.4) 42.0 | 41.8 (19.2) 39.5 | 48.6 (25.6) 46.5 | 45.2 (16.2) 45.0 | 44.0 (13.1) 45.0 |
| **Age at Baseline (years) distribution, n (%)** | | | | | | | |
| **18-24** | 135 (10.7) | 80 (10.0) | 52 (11.7) | 24 (26.7) | 3 (25.0) | 104 (11.7) | 7 (7.5) |
| **25-34** | 256 (20.3) | 148 (18.5) | 107 (24.0) | 19 (21.1) | 1 (8.3) | 167 (18.7) | 18 (19.4) |
| **35-44** | 243 (19.3) | 157 (19.6) | 84 (18.8) | 7 (7.8) | 2 (16.7) | 164 (18.4) | 21 (22.6) |
| **45-54** | 239 (19.0) | 153 (19.1) | 85 (19.1) | 13 (14.4) | 1 (8.3) | 166 (18.6) | 26 (28.0) |
| **55-64** | 220 (17.5) | 167 (20.9) | 52 (11.7) | 15 (16.7) | 1 (8.3) | 182 (20.4) | 17 (18.3) |
| **65-74** | 110 (8.7) | 71 (8.9) | 37 (8.3) | 7 (7.8) | 2 (16.7) | 78 (8.8) | 3 (3.2) |
| **≥ 75** | 56 (4.5) | 25 (3.1) | 29 (6.5) | 5 (5.6) | 2 (16.7) | 30 (3.4) | 1 (1.1) |
| **Total** | 1259 | 801 | 446 | 90 | 12 | 891 | 93 |
| **P-value (Baseline v Analysis Sample - Sex)** | | 0.001† | | | | | |

* All participants ≥ 18, which were included in baseline study; ** participants, which participated in both studies; # participants, which did not come back to the follow-up study; $ participants, which entered the study at follow-up study, analysed at follow-up time point, § Sub-population used for T cell analysis in follow-up study; [†] Fisher's exact

**Supplementary Table 2. Change in median IgG antibody titers from baseline to follow-up**

| | n | Baseline (Median (IQR)) | Follow-up (Median (IQR)) | Wilcoxon signed rank p-value | %decrease in median |
|---|---|---|---|---|---|
| **Euroimmun** | 801 | 0.6 (0.2-3.6) | 0.3 (0.1-1.3) | <0.000 | 50.0 |
| **Abbott** | 801 | 0.9 (0.0-5.8) | 0.1 (0.0-0.7) | <0.000 | 88.9 |
| **Roche** | 40 | 29.0 (8.9-50.4) | 12.0 (4.0-35.0) | <0.000 | 58.6 |

IQR = interquartile range

**Supplementary Table 3. Seroprevalence**

| Sample | Baseline (Adults)* | Baseline Analysis Sample (Adults)** | Follow-up Analysis Sample[#] | New at Follow-up[$] | All Adults at Follow-up |
|---|---|---|---|---|---|
| % positive (number positive/total number) | 45.0 (566/1259) | 51.4 (412/801) | 45.4 (364/801) | 37.8 (34/90) | 44.7 (398/891) |
| 95% Confidence Interval | 42.2 - 47.8 | 47.9-54.9 | 42.0 - 49.0 | 27.6 - 48.6 | 41.4 - 48.0 |

* All participants ≥ 18, which were included in baseline study, analysed at baseline time point; ** participants, which participated in both studies, analysed at baseline time point; # participants, which participated in both studies, analysed at follow-up time point; $ participants, which entered the study at follow-up time, analysed at follow-up time point

**Supplementary Table 4: Potential new infections between baseline and follow-up**

| Sex | Age+ | Antibody baseline (I1) | | | Antibody follow-up (I2) | | | Serostatus$ | positive PCR between I1 and I2# | Symptoms between I1 and I2# |
|---|---|---|---|---|---|---|---|---|---|---|
| | | Anti S IgG (OD ratio)* | Anti N IgG (RLU)** | Neutralization assay § | Anti S IgG (OD ratio)* | Anti N IgG (RLU)** | Neutralization assay § | | | |
| male | 25-34 | 0.195 | 0.0 | ≤1:4 | 2.830 | 5.8 | 1:256 | n-p | - | Cough |
| male | 45-54 | 0.325 | 0.1 | ≤1:4 | 4.149 | 7.3 | 1:16 | n-p | - | Fever, headache, irritated eyes, sore throat, running nose |
| female | 55-64 | 0.203 | 0.0 | ≤1:4 | 2.662 | 0.7 | 1:64 | n-d | - | Sleep disturbance |
| male | 25-34 | 0.075 | 0.0 | ≤1:4 | 1.638 | 2.0 | 1:64 | n-p | + 10/2020 | Cold, sore throat, running nose |

+ at baseline; * OD ratio >0.8= positive ; ** RLU >1.4 positive; §cutoff: >1:4=positive, ≤1:4=negative; # self-reported; $ p = positive, d = discrepant, a = IgA only, n = negative
OD= optical density. RLU= relative light unit. S= spike protein. N= nucleocapsid protein

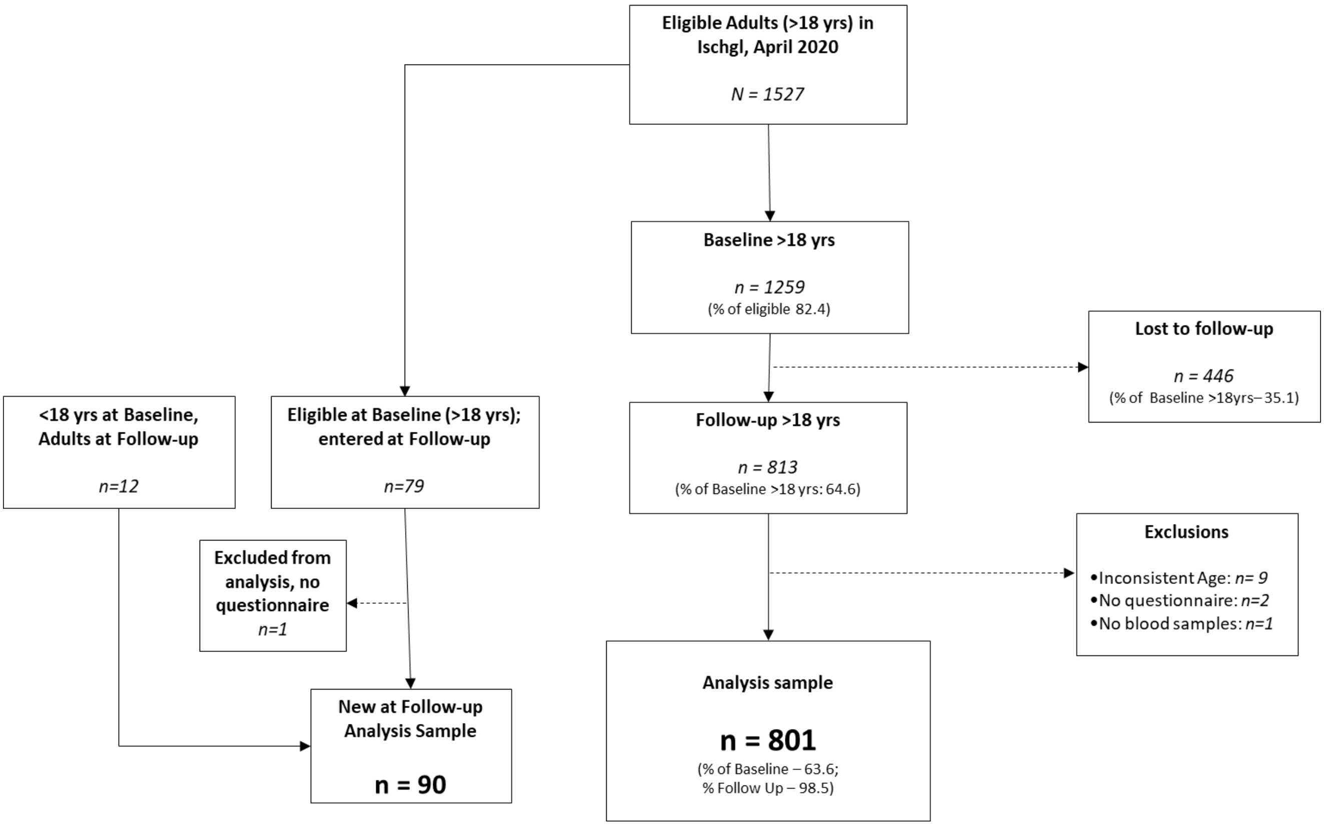

**Supplementary Figure 1. Study enrolment follow-up (Ischgl 2)**

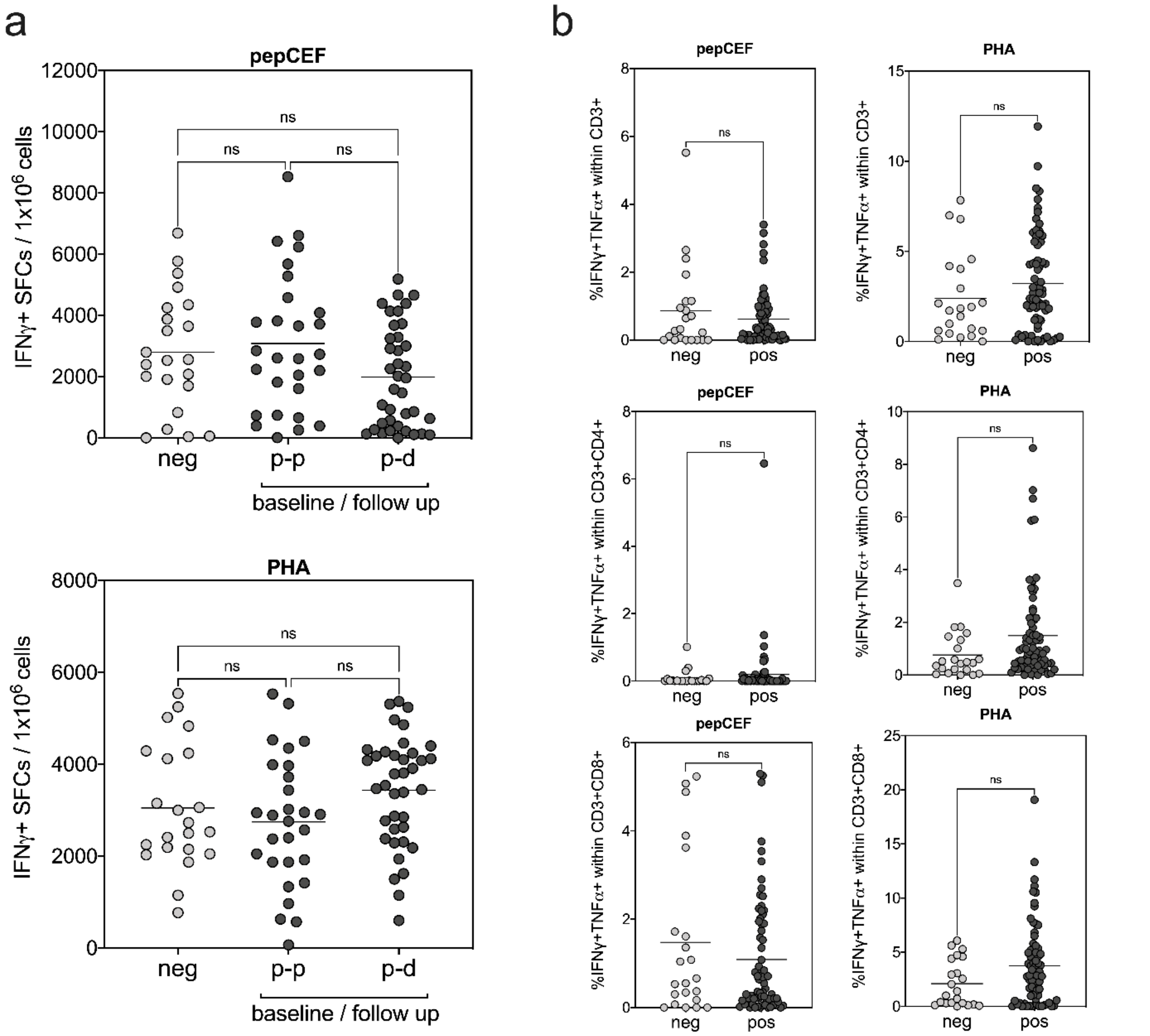

**Supplementary Figure 2. T cell responses against positive controls. (a)** IFNγ ELISPOT assay and **(b)** IFNγ/TNFα intracellular cytokine (IC) staining after a 7-day *in vitro* expansion followed by re-stimulation with pepCEF peptide pool and PHA as positive controls. Values after stimulation were normalized to non-stimulated samples in both ELISPOT and IC stainings. **(a)** Shown are IFNγ positive SFCs per $10^6$ cells in baseline negative and positive study groups (serostatus at follow-up study either positive p or discordant d) after stimulation with pepCEF (upper graph) or PHA (lower graph). **(b)** Percentage of IFNγ/TNFα-producing CD3+ total T cells, CD3+CD4+ helper T cells and CD3+CD8+ cytotoxic T cells after are shown. After IC staining living/singlet cells were gated for CD3+ total, CD3+CD4+ and CD3+CD8+ T cells and the indicated percentage depicts the frequency of IFNγ/TNFα double-positive cells in the respective population of the sample stimulated with pepCEF or PHA minus the frequency of the non-stimulated control.
